# Bridging medicine access gaps through cross-national market linkage in South Korea and Italy: a conceptual case

**DOI:** 10.64898/2026.10.08.26365076

**Authors:** Minji Sohn, Umberto M. Musazzi

**Affiliations:** College of Pharmacy, Sahmyook University, 815 Hwarang-ro, Nowon-gu, Seoul, Republic of Korea; Department of Pharmaceutical Sciences, University of Milan, Milan, Italy

**Keywords:** Medicine shortages, Pharmaceutical policy, Reimbursement, Drug supply, Essential medicines, Republic of Korea, Italy, Market Access

## Abstract

**Background:** Medicine shortages are monitored within national borders, but complementarity across national markets is poorly characterised.

**Objective:** To characterise medicines supplied through South Korea’s national access programme and to quantify this complementarity between South Korea and Italy.

**Methods:** In this cross-sectional study, we linked shortage records from the Italian Medicines Agency and Korean Ministry of Food and Drug Safety, Korea Orphan and Essential Drug Center supply notices, and national reimbursement lists. The primary outcome was the proportion of shortage configurations with a reimbursed counterpart in the partner country, identified by normalised ingredient, pharmaceutical form, and route, and not recorded as withdrawn or in shortage. The primary analysis covered Jan 1–June 30, 2026, and the secondary analysis covered 2023–26.

**Results:** Among 341 formulations supplied through the Korean national access programme, 242 (71.0%) lacked domestic marketing authorisation at first observed supply or were supplied after market withdrawal. Of 338 Italian shortage configurations in the first half of 2026, 228 (67.5%) had an available reimbursed Korean counterpart; over 2023–26, 332 of 549 (60.5%) did. Conversely, 27 of 47 Korean configurations (57.4%) had such an Italian counterpart; over 2023– 26, 93 of 184 (50.5%) did. In both directions, ingredients on EU critical or Korean essential medicine lists more often had a cross-border match than unlisted ingredients.

**Conclusions:** About half of shortage or access-gap medicines had a potentially available reimbursed counterpart in the other country. Cross-national linkage of medicine data may help identify potential counterparts, although these require regulatory, clinical, and procurement review.

**Research in context:** *What is already known about the topic?:* Studies have mainly compared shortage patterns or management across countries or described importation and information sharing as responses. A 2025 US analysis found that, for about two-thirds of generic sterile injectables in shortage with three or fewer US manufacturers, a version was approved in peer countries. We found no study that linked national shortage records to a partner country’s reimbursement market while accounting for withdrawals and concurrent shortages.

*What does this study add to the literature?:* In the first half of 2026, about two-thirds of Italian and more than half of Korean shortage or access-gap configurations had an available reimbursed counterpart abroad, and critical or essential medicines more often had a cross-border match. Most medicines supplied through South Korea’s national access programme lacked domestic marketing authorisation at first supply or were supplied after market withdrawal.

*What are the policy implications?:* Linking national registers can identify candidate counterparts across borders, but each requires product-level regulatory and procurement review and an institution able to act on the resulting information. Wider application would require standardised reporting of ingredient, form, route, and shortage date.

## Background

National health systems rely on market mechanisms, regulatory approval pathways, and designation systems to secure patient access to medicines [1]. Since the World Health Organization (WHO) introduced the essential medicines concept in 1977, countries have built instruments to prioritise medicines addressing major population health needs [2,3], and the European Union added the Union List of Critical Medicines in December 2023 for products at risk of supply disruption [4]. Medicines nonetheless remain unavailable through routine domestic channels because they lack domestic approval, have been withdrawn after commercial non-viability, or face demand the domestic market cannot absorb [5–8]. During a national shortage of cytarabine, patients with acute myeloid leukemia were 47% less likely to begin induction chemotherapy within 14 days of diagnosis [9].

Designation systems classify medicines by therapeutic importance, rarity, or anticipated risk, not by observed access barriers, so medicines facing real-world access problems may fall outside them [10,11]. Shortage management remains national or supranational (eg, EU) in scope, with little systematic exchange of supply information across borders [12], even though manufacturing failures, commercial withdrawals, and regulatory non-approvals may affect multiple countries [13,14].

We examined whether medicines recorded as unavailable in one country could be identified in the routine reimbursement market of a partner country. South Korea and Italy both publish national medicine registers: Korea through the Ministry of Food and Drug Safety (MFDS) shortage register and the Korea Orphan and Essential Drug Center (KOEDC), and Italy through the Agenzia Italiana del Farmaco (AIFA). We quantified bidirectional complementarity and explored whether counterpart status was associated with clinical importance.

## Methods

### Study design and data sources

This cross-national study used publicly available national medicine datasets from South Korea and Italy. The primary matching analysis was cross-sectional, with Korean programme activity additionally described over 21 quarters. Because only publicly available government data were used, institutional review board approval was not required. Reporting followed STROBE [15].

Italian shortage data were obtained from the AIFA Medicinali Carenti database (version updated July 21, 2026; accessed July 27, 2026). Records whose stated reason contained cessata commercializzazione or sospensione commercializzazione were classified as marketing withdrawals. After their exclusion, 1,956 shortage records remained, representing 323 unique active ingredients in the first half of 2026 and 563 unique active ingredients from 2023 to 2026. Korean shortage data came from MFDS reports (January 2023 to July 2026) and KOEDC quarterly supply notices covering January 2021 through March 2026 (21 quarters). Routine markets were represented by the Health Insurance Review and Assessment Service (HIRA) reimbursement database (June 2026; 1,627 unique ingredients after name normalisation) and the AIFA Classe A and Classe H lists (January 31, 2026; 1,323 unique ingredients). Additional details on the data sources are provided in **Table A.1**, and the classification of each list and record type in **Table A.2**.

The primary window was January 1 to June 30, 2026, with a secondary analysis over 2023 to 2026. AIFA shortages starting after the snapshots, which were notified before onset, were excluded. Cessations notified under Article 23a of Directive 2001/83/EC were treated as marketing withdrawals (**Appendix A.1**) [16]. The Korean register records the reporting date instead of the onset and does not flag advance notifications.

### Shortage and access-gap definitions

The Korean shortage register captures only specified categories of finished product and is otherwise voluntary, so it under-represents domestic unavailability (Box 1). We therefore defined the Korean denominator as medicines authorised in Korea that were nevertheless not supplied through the routine market: reported shortages together with KOEDC-supplied medicines holding a domestic marketing authorisation. Formulations not authorised in Korea by the quarter of first supply (174 of 341), those whose authorisation status could not be determined (23), or those whose domestic authorisation had been withdrawn before the first observed supply or for which withdrawal date was not available (62) were excluded; six formulations whose authorisation was withdrawn during the observation window after supply had begun were retained (**Appendix A.2**). Programme formulations and shortage records were subjected to prespecified category exclusions, removing products that could not be meaningfully matched across national reimbursement lists (**Appendix A.2**). Marketing withdrawals were distinguished from active shortages in both registers.

Manufacturing source or therapeutic modality was not itself a criterion for exclusion. Medicines were retained when a standardised identity and route of administration could be defined and compared across countries. We excluded products for which meaningful cross-national comparison was precluded by product configuration, intended use, or insufficiently standardised composition, including integrated procedural products, biomaterials, ex vivo organ-preservation solutions, instrument disinfectants, heterogeneous allergen and herbal preparations, and probiotics. Diagnostic agents, radiopharmaceuticals, and routine hydration, electrolyte, diluent, dialysis, or nutrition solutions were also excluded. Prophylactic vaccines were excluded because their separate public-health procurement and distribution systems could result in incomplete ascertainment in the routine reimbursement data used for counterpart identification; they were therefore treated as not evaluable (14 Italian and 24 Korean register records, and 34 designated formulations). For cell therapy products, a standardised ingredient identity cannot be established without standardised nomenclature because of process-dependent biological heterogeneity. We therefore included cell therapies with an assigned WHO International Nonproprietary Name (INN; eg, tisagenlecleucel) and excluded those without an INN from ingredient-level matching.

### Matching

Ingredient names were resolved to normalised ingredient entities using a single rule set applied to both countries, defined by clinical distinctness rather than chemical structure alone. Salt, hydrate, and solvate modifiers were collapsed to the parent ingredient, whereas derivatives with clinically meaningful differences in dose, route of administration, bioavailability, or duration of action were retained as separate entities. These included prodrug esters, topical and depot esters, and single enantiomers. Liposomal formulations were likewise kept separate from conventional formulations of the same ingredient (**Appendix A.2**). Truncated or fragmented ingredient names resulting from source parsing were traced to the original public records and reconstructed before the prespecified rules were applied. Matching required concordance of the normalised ingredient entity (hereafter ingredient), pharmaceutical form (hereafter form), and route of administration, the latter two coded to European Directorate for the Quality of Medicines and HealthCare (EDQM) standard terms (**Appendix A.2**). In the designation lists, oral ingredients for which a single pharmaceutical form could be established from unambiguous registry or originator-label evidence were assigned to that specific form. Entries for which multiple forms remained plausible retained their route classification but were considered indeterminate at the formulation level and excluded from formulation-level matching. The criticality and designation lists are published at the parent-substance level, so concordance with those lists was assessed after collapsing ingredients to their active moiety. A sensitivity analysis treated the various immediate-release oral forms as one and the same pharmaceutical form, following the definition of “generic” medicines reported in Article 10(2)(b) of Directive 2001/83/EC. Strength was not part of the matching key, and fixed-dose combinations were matched as complete combinations.

A counterpart was counted as available when at least one reimbursed product with the same ingredient, form, and route remained after excluding products withdrawn or discontinued before June 30, 2026, as well as products reported in shortage in the same analysis window. Two sensitivity analyses varied this definition (**Appendix A.2**).

### Outcomes

The primary outcome was the proportion of shortage or access-gap medicines with an available reimbursed counterpart in the partner country. Secondary outcomes included counterpart identification based on reimbursement listing alone and representation of medicines with and without counterparts on the EU Union List of Critical Medicines or the Korean national essential medicine list. The KOEDC programme was additionally characterised by domestic regulatory status, orphan-drug and national essential-medicine designation status, Medical Dictionary for Regulatory Activities (MedDRA) System Organ Class (SOC) based on the primary indication, and longitudinal supply activity.

## Statistical analysis

Categorical variables were reported as counts and percentages. Results are reported descriptively as counts and proportions of all records in the registers for each period. Proportions were compared using two-sided Fisher exact tests. Comparisons against the criticality and essential-medicine lists were exploratory and not prespecified as a fixed set; therefore, nominal p values were reported without adjustment for multiplicity and should not be interpreted as familywise inference. Longitudinal changes in programme activity over time were evaluated using the Mann–Kendall trend test. Sensitivity analyses that removed the category exclusions, added Italian temporary cessations of marketing to the shortage denominator, and counted programme formulations with undetermined authorisation as authorised are reported in **Table A.3**. Analyses used R version 4.4.1 and Python version 3.12.

## Results

### Characteristics of programme-supplied medicines

Between January 2021 and March 2026, 341 formulation-level entries corresponding to 235 unique active ingredients appeared in KOEDC quarterly notices. Of these, 174 (51.0%) had no domestic marketing authorisation by the quarter in which supply was first observed and a further 68 (19.9%) were supplied after market withdrawal, so 242 (71.0%) fell outside conventional approved-market pathways; authorisation status at first supply could not be determined for a further 23 (6.7%) (**Figure 1**). Seventy-five (22.0%) carried orphan-drug designation and 167 (49.0%) national essential-medicine designation, and 128 (37.5%) neither; that undesignated group had the lowest overlap with the EU Union List of Critical Medicines (12.5%, vs 24.9% overall). The therapeutic-area distribution of the portfolio is shown in **Fig. A.1**.

**Figure 1.**
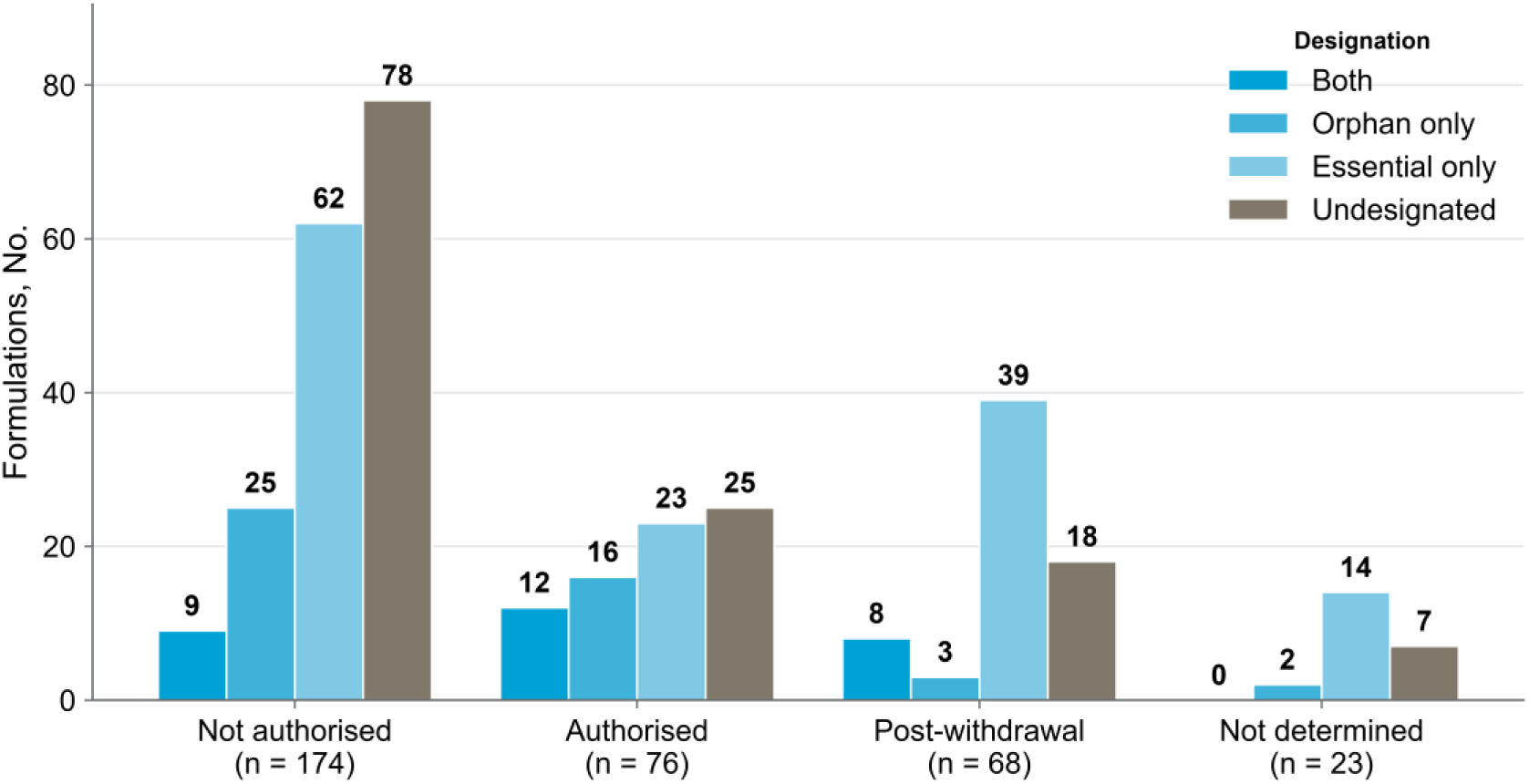
Regulatory Status of Medicines Supplied Through Korea’s National Access Programme. Formulations are grouped by domestic regulatory status at the time of first observed supply and stratified by orphan-drug and essential-medicine designation status. Formulations were supplied through the Korea Orphan and Essential Drug Center, January 2021 to March 2026 (N = 341). Designation categories are mutually exclusive as shown, and orphan and national essential-medicine designations were assessed at the active-ingredient level.

Programme activity intensified over the period. Actively supplied products rose from 101 to 146 (+44.6%), a non-significant trend (Mann–Kendall τ=0.251; p=0.12), whereas quarterly transactions rose by 95.4% (τ=0.819), pack volume by 134.1% (τ=0.743), and dosage units by 181.9% (τ=0.800; all p<0.0001) (**Fig. A.2**).

### Cross-national counterpart identification

Of the 338 combinations of ingredient, form, and route in shortage in Italy in the first half of 2026, 228 (67.5%) had an available Korean reimbursed counterpart with the same form and route; over 2023–26, 332 of 549 (60.5%) had such a counterpart. In the reverse direction, 27 of 47 Korean combinations (57.4%) had an available Italian counterpart in the first half of 2026 and 93 of 184 (50.5%) over 2023–26 (**Table 1** and **Figure 2**). When supply status was not considered, the corresponding proportions were 235 of 338 (69.5%) and 351 of 549 (63.9%) in the Italian direction and 29 of 47 (61.7%) and 97 of 184 (52.7%) in the Korean direction (**Table A.3**).

**Figure 2.**
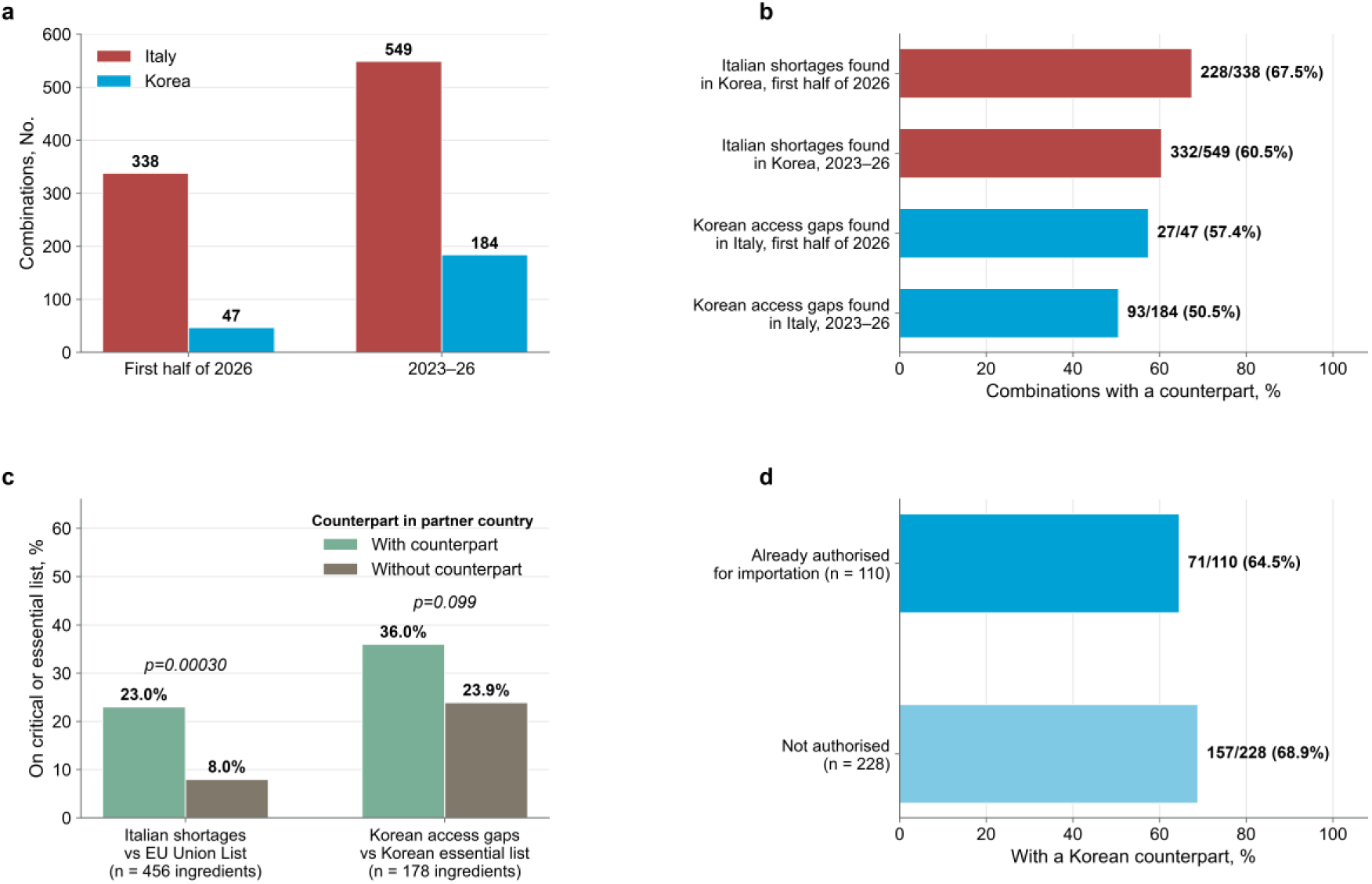
Cross-National Complementarity of Medicine Shortages Between Korea and Italy. (a) Ingredient, form, and route combinations included in each analysis window. (b) Proportions with a reimbursed counterpart in the partner country, by direction and window. (c) Critical or essential medicine designation among ingredients with and without a counterpart in the partner country (2023–26). (d) Counterpart proportions among Italian shortages with and without authorisation for importation. Matching required concordance of normalised ingredient, EDQM pharmaceutical form, and route of administration.

**Table 1.** Bidirectional Cross-National Counterpart Identification Between South Korea and Italy.

| **Direction and Window** | **Ingredients, No.** | **Matched by Ingredient, No. (%)** | **Ingredient, Form, and Route Combinations, No.** | **Matched by Ingredient, Form, and Route, No. (%)** | **On EU Critical Medicines List, No. (%)** |
| --- | --- | --- | --- | --- | --- |
| Italy shortage to Korea, first half of 2026 | 304 | 248 (81.6) | 338 | 228 (67.5) | 55 (18.1) |
| Italy shortage to Korea, 2023–26 | 456 | 344 (75.4) | 549 | 332 (60.5) | 88 (19.3) |
| Korea shortage or program to Italy, first half of 2026 | 47 | 29 (61.7) | 47 | 27 (57.4) | NA |
| Korea shortage or program to Italy, 2023–26 | 178 | 111 (62.4) | 184 | 93 (50.5) | NA |
| MFDS-designated access gap to Italy | 333 | 138 (41.4) | NA | 115 (34.5) | NA |
| Italian shortages authorised for importation | 92 | 73 (79.3) | 110 | 71 (64.5) | NA |
| Italian shortages not authorised for importation | 212 | 175 (82.5) | 228 | 157 (68.9) | NA |

Abbreviations: EDQM, European Directorate for the Quality of Medicines and HealthCare; MFDS, Ministry of Food and Drug Safety; NA, not applicable. Matching required concordance of the normalised ingredient entity, pharmaceutical form, and route of administration (the latter two coded to EDQM standard terms), with salt, hydrate, and solvate modifiers collapsed to the parent ingredient and clinically differing derivatives kept separate. Strength was not matched. The Korean denominator comprises reported shortages and Korea Orphan and Essential Drug Center-supplied medicines holding a domestic marketing authorisation; the two sources shared no ingredient in the first half of 2026 and 11 of 40 (27.5%) over 2023–2026. The 2 directions are not directly comparable because shortage reporting is category-specific in Korea but universal in Italy.

Of the 110 Italian combinations without an available counterpart in the first half of 2026, 52 involved an ingredient absent from the Korean reimbursement list, 51 an ingredient present only in a different form or route, and seven a listed counterpart whose products were all withdrawn or in shortage. The corresponding numbers in the Korean direction were 17, one, and two. Treating the various immediate-release oral forms as one pharmaceutical form raised the primary estimates to 75.4% and 68.4% in the Italian direction and 57.4% and 55.7% in the Korean direction (**Table A.3**).

Italian shortages already authorised for importation matched the Korean reimbursement list at a rate no higher than those not authorised (73 of 92 [79.3%] vs 175 of 212 [82.5%] by ingredient; 71 of 110 [64.5%] vs 157 of 228 [68.9%] by ingredient, form, and route). In a separate analysis of MFDS-designated access-gap medicines, 791 of 903 designated formulations remained after prespecified exclusions. Of these, 435 were themselves reimbursed in Korea, leaving 356 designated access-gap formulations. The 356 formulations represented 333 unique ingredients, of which 115 (34.5%) had an Italian counterpart matching on ingredient, form, and route and 138 (41.4%) matched on ingredient alone. The corresponding same-form-and-route proportions were 30 of 148 (20.3%) for the essential subgroup and 89 of 196 (45.4%) for the orphan subgroup.

### Clinical importance of matched medicines

The EU Union List of Critical Medicines resolved to 282 active ingredients and the Korean national essential medicine list to 395, of which 130 were common to both (**Fig. A.3**). Among the 304 Italian shortage ingredients in the first half of 2026, 55 (18.1%) appeared on the EU list and 55 (18.1%) on the Korean list; over 2023–26, 88 of 456 (19.3%) appeared on the EU list and 86 (18.9%) on the Korean list. Counterpart availability was compared between ingredients on and off each list, in both directions and both windows, giving eight exploratory comparisons reported in full without adjustment for multiplicity (**Table A.4**). Listed ingredients more often had a counterpart in all eight comparisons, and three reached nominal significance, all among Italian shortage ingredients: on the EU list in the first half of 2026 (53 of 55 [96.4%] vs 195 of 249 [78.3%]; p=0.00087) and over 2023–26 (79 of 88 [89.8%] vs 265 of 368 [72.0%]; p=0.00030), and on the Korean essential list over 2023–26 (76 of 86 [88.4%] vs 268 of 370 [72.4%]; p=0.0014). For the remaining five, p values ranged from 0.099 to 0.37.

Across the two sources of the Korean denominator, national essential designation was carried by 44.8% of ingredients drawn from the national access programme and 26.8% of those drawn from shortage reports (13 of 29 vs 37 of 138; p=0.073; the remaining 11 ingredients appeared in both sources, six of which were designated), and 56 of the 178 Korean ingredients (31.5%) carried it before any matching was performed, compared with 86 of 456 (18.9%) on the Italian side.

Orphan designation followed the opposite pattern. Almost no orphan-designated medicine appeared in the Italian shortage list: two of the 304 ingredients in the first half of 2026 (0.7%) and three of 456 (0.7%) over 2023–26 were on the AIFA orphan-medicine list, whereas 75 of 341 Korean programme formulations (22.0%) carried orphan designation.

### Market withdrawals

Between 2023 and 2026, 317 active ingredients were reported as discontinued in Korea and 157 in Italy (**Table A.5**). Of the Korean discontinuations, 149 (47.0%) remained listed in Italy with at least one product not withdrawn there; of the Italian withdrawals, 107 (68.2%) remained listed in Korea. Twenty-seven ingredients had withdrawal records in both countries (**Table A.6**). The proportion designated critical was 29.6% among ingredients discontinued in both countries and 14.5% among those discontinued in Korea alone (8 of 27 vs 42 of 290; p=0.052) (**Fig. A.4**).

## Discussion

In this cross-national study, more than half of medicines unavailable in one country had a counterpart on the partner country’s reimbursement list, in both directions. The directional proportions were not directly compared because the national reporting systems differ in coverage. The medicines with potential counterparts were not confined to those of lower clinical importance: ingredients on a critical or essential medicine list more often had a counterpart in all eight exploratory comparisons, with three reaching nominal significance, all among Italian shortages (**Table A.4**). The two national criticality frameworks also agreed on fewer than half of what either designates.

Our findings are broadly concordant with the closest comparable analysis we identified. Of 81 generic sterile injectables in shortage in the USA between 2021 and 2024 that had three or fewer US manufacturers approved by the US Food and Drug Administration (FDA), 53 had at least one manufacturer with a version approved in one or more of six peer countries but not by the FDA [24]. Whereas that one-directional analysis assessed regulatory approval of sterile injectables, the present study examined the included medicine types in both directions and required counterparts to be reimbursed and not recorded as withdrawn or in shortage.

A recent analysis of systemic antibacterials in Europe found that shortages clustered by product characteristics (older, off-patent, parenterally administered agents) rather than by country [25], implying that national lists record overlapping manifestations of shared upstream problems.

More than a third of the portfolio, 128 of 341 formulations (37.5%), carried neither orphan-drug nor national essential-medicine designation, and this group had the lowest overlap with the EU Union List of any designation category (12.5%, vs 24.9% overall), even though routine domestic access had already failed for each of them. Hungary showed the same divergence: only 190 of 605 shortage medicines were on its critical list [26]. Designation systems anticipate supply risk or recognise therapeutic priority rather than detect access that has already lapsed: essential-list selection weighs disease burden, benefit, and cost-effectiveness without regard to commercial availability [3], and orphan designation follows prevalence [27].

For 68 formulations (19.9%), the programme continued to supply a medicine after its domestic authorisation had lapsed, a pattern that studies of low-margin generic and sterile injectable products have attributed to commercial rather than clinical decisions [6–8]. The Italian regulatory framework similarly includes unavailability of medicines on the market caused by temporary or permanent voluntary market withdrawal within carenze, while distinguishing them from geographically localised distribution failures [28]. Commercial exit is not the only route to access failure: manufacturing-quality failures at generic producers have also removed products for extended periods and propagated shortages across therapeutic classes [29].

The two systems authorise supply from outside the routine market through structurally different routes. Italy permits healthcare facilities to import medicines without national authorisation for specific patients, under the responsibility of a physician and subject to prior authorisation by AIFA [30]. A similar but separate procedure exists for authorised medicines that are temporarily unavailable [19]; the records analysed here correspond to the latter, for which requests originate from individual healthcare facilities [28]. Korea channels equivalent requests through a single national body that can supply without any shortage report, and Australia places the coordinating function in the regulator instead [31]. Although AIFA governs and oversees the importation of medicinal products into Italy, there is no Italian national counterpart to the KOEDC as a coordinating procurement structure, an institutional asymmetry relevant to how cross-national linkage could be acted on. Procurement design may itself affect supply resilience: concentrated tendering can reduce supplier diversity [32], while pooled or centralised arrangements depend on stable institutional coordination, demand forecasting, and purchasing commitments [33,34].

Linking the two registers identifies medicines recorded as unavailable in one country but routinely reimbursed in the other. Each potential counterpart requires product-level review before it can be acted on. These findings support a mutual medicine access system in which each country’s shortage or access-gap records are routinely linked to the other’s reimbursement database, generating candidate lists for expert regulatory and procurement review (**Figure 3**). Standardised reporting of ingredient, pharmaceutical form, route, and date would facilitate routine cross-national linkage. Cross-national linkage would supplement domestic shortage responses such as substitution.

**Figure 3.**
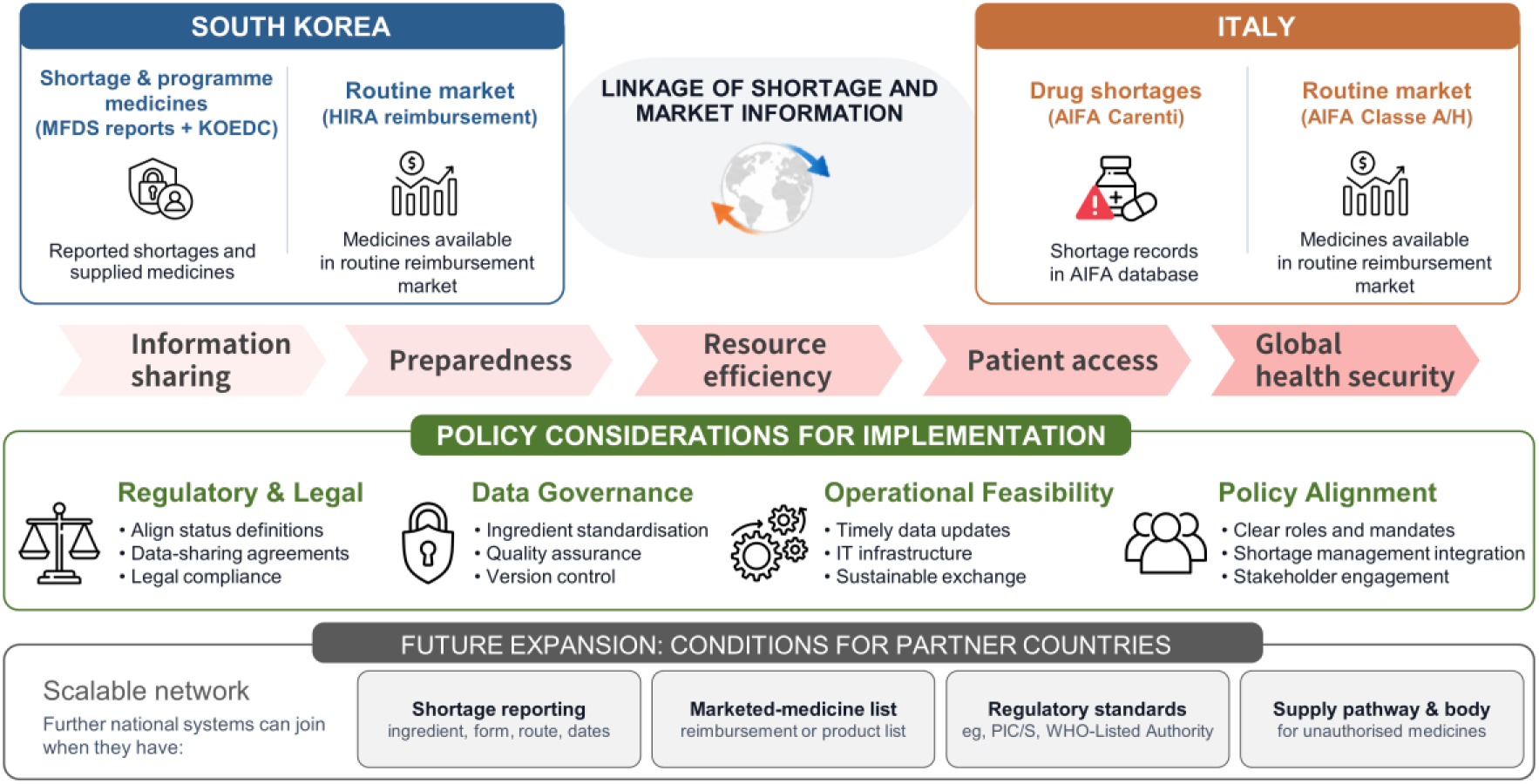
Framework for Cross-National Medicine Exchange and Policy Implementation. Bidirectional linkage of national shortage or access-gap records with partner-country market information to identify potential counterparts. Abbreviations: AIFA, Italian Medicines Agency; HIRA, Health Insurance Review and Assessment Service; KOEDC, Korea Orphan and Essential Drug Center; MFDS, Ministry of Food and Drug Safety; PIC/S, Pharmaceutical Inspection Co-operation Scheme.

On the Korean side the receiving institution already exists in statute. The Pharmaceutical Affairs Act places a standing council on the stable supply of national essential medicines within MFDS and establishes the KOEDC. Amendments taking effect on November 12, 2026, will add powers to commission production and directly import national essential medicines, both of which will be delegated to the KOEDC [20].

The European Union has moved in this direction through the reform of the EU pharmaceutical legislation (so-called EU pharma package) and the proposed Critical Medicines Act, which envisage voluntary joint procurement initiated at a member state’s request [35,36]. A comparable information infrastructure is already being assembled within the Union: the regulation reinforcing the European Medicines Agency’s crisis role created a steering group on shortages and a European Shortages Monitoring Platform [17]. Even in more decentralised systems, cross-national data linkage could help identify alternative sources during shortages.

Operational barriers may remain even where potential counterparts are available. Cross-border transfer requires resolution of labelling and language requirements, national batch release, and mutual recognition of manufacturing standards, and a partner under its own supply pressure may restrict export [37]. The European Union has concluded mutual recognition agreements on good manufacturing practice with seven third countries, but Korea is not among them [38]. Both participate in the Pharmaceutical Inspection Co-operation Scheme [39], and MFDS is a WHO-Listed Authority across all regulatory functions [40]. Cross-national redistribution also raises an allocation question: improving access in one country should not worsen a shortage in the supplying country, and any procurement mechanism would therefore require safeguards for domestic supply, product quality, and equitable allocation [5].

A cross-national match indicates that a corresponding medicine is present on the partner country’s reimbursement list and not recorded as withdrawn nor in shortage. It does not establish therapeutic equivalence, regulatory interchangeability, commercial availability, or exportable stock, and every match requires product-level review. Parenteral presentations were matched at a single coarse level because the two registers record different attributes: the Korean reimbursement list identifies a product as an injection, whereas the Italian list provides administration route but incompletely records pharmaceutical form. This may create counterparts that differ in practice. The two reference lists also differ in breadth, further limiting comparison of the directional match rates. The registers differ in both coverage and timing. The Italian register records shortage onset, whereas the Korean register records the reporting date. Korean reporting is mandatory only for designated categories, whereas the Italian duty attaches to every national marketing authorisation, so shortages of non-designated Korean medicines are likely under-captured. Both reference lists are reimbursement lists rather than measures of active supply. Although counterparts recorded as withdrawn or shortage were excluded, products that remain authorised without such reports but are not actively supplied cannot be identified. Counterpart rates may therefore overestimate immediately available supply. The design also cannot distinguish the upstream causes of shortage, so a match identifies only a candidate alternative source. Finally, the findings reflect two high-income systems and may not generalise to countries with different regulatory, reimbursement, or procurement structures.

## Conclusions

In both directions, about half of shortage or access-gap medicines had an available reimbursed counterpart in the partner country. Routinely linking existing national registers could generate candidate lists for regulatory and procurement review, provided each country has an institution able to act on them.

## Ethics approval

The study analyzed aggregate, non-identifiable administrative data that are publicly available from national agencies. It did not involve human participants or identifiable personal data; institutional review board approval and informed consent were therefore not required.

## Data statement

All primary datasets are publicly available from the agency sources cited. Shortage and national access programme data used for the primary analyses are provided in Appendix B.

**Appendix A.** Methods A.1–A.2, Tables A.1–A.6, Figs. A.1–A.4

**Appendix B.** Supplementary data

#### Box 1. Regulatory Definitions of Medicine Shortage and Market Withdrawal

**European Union**. Regulation (EU) 2022/123, Article 2, defines a shortage as “a situation in which the supply of a medicinal product that is authorised and placed on the market in a Member State or of a CE-marked medical device does not meet demand for that medicinal product or medical device at a national level, whatever the cause” [17]. Cessation of marketing is governed separately: under Article 23a of Directive 2001/83/EC, the marketing authorization holder must notify the competent authority of temporary or permanent cessation, in principle no less than 2 months in advance [16].

**Italy.** Under the AIFA reporting framework, a medicine is listed as carente (in shortage) when the marketing authorization holder notifies a temporary interruption of supply. The same public list also records cessata commercializzazione and sospensione commercializzazione, allowing temporary or permanent cessation of marketing to be distinguished from other reasons for shortage. Under Article 34(6) of Legislative Decree 219/2006, as amended in December 2025, the marketing authorization holder must notify AIFA of temporary or permanent cessation of marketing at least 2 months in advance [18]. Temporary shortages are handled separately, including through authorization to import the medicine from abroad [19].

**Korea.** Reporting obligations under the Pharmaceutical Affairs Act and MFDS Notification No. 2025-33 apply to finished pharmaceutical products designated by the Minister of Health and Welfare in consultation with the Minister of Food and Drug Safety [20–23]. Marketing authorization holders and importers must report intended discontinuations at least 180 days in advance (or within 10 days after discontinuation in unavoidable cases), and report within 1 month any planned reductions expected to result in suspension of production or importation for 3 months or more. Voluntary reporting is also permitted for holders not subject to these requirements. Consequently, shortages of non-designated medicines may be less comprehensively captured.

## Data Availability

All data produced in the present study are available upon reasonable request to the authors

## Acknowledgements

Haeun Cho (College of Pharmacy, Sahmyook University), a pharmacy student, assisted with the verification of Korea Orphan and Essential Drug Center records. She provided written permission to be named.

## CRediT authorship contribution statement

Minji Sohn: Conceptualization, Methodology, Data curation, Investigation, Formal analysis, Validation, Writing – original draft, Supervision. Umberto M. Musazzi: Conceptualization, Methodology, Data curation, Investigation, Formal analysis, Validation, Writing – review & editing. Both authors accessed and verified the underlying data and approved the final manuscript.

